# Left ventricular function and its longitudinal change in Swiss childhood cancer survivors – Results from the CardioOnco Study

**DOI:** 10.1101/2025.11.20.25340636

**Authors:** Severin Fankhauser, Yara Shoman, Fabiën N Belle, Eva Hägler-Laube, Moritz J Hundertmark, Gabriela M Kuster, Eva Scheler, Tomáš Sláma, Nicolas von der Weid, Claudia E Kuehni, Reto D Kurmann, Christina Schindera

## Abstract

**Background/Purpose:** Childhood cancer survivors (CCS) are at risk for cardiac late effects. Echocardiographic assessment of cardiac function by left ventricular ejection fraction (LVEF) is recommended for survivors treated with known cardiotoxic treatments (anthracyclines; heart-relevant radiotherapy). The evidence on cardiotoxicity of other systemic anticancer therapies is conflicting and longitudinal changes in heart function are understudied. We assessed cardiac function in CCS; factors associated with LVEF and longitudinal changes in LVEF.

**Methods:** In this prospective, multicenter cohort study, we invited CCS aged ≥18 years, diagnosed <21 years, survived ≥5 years, and received any systemic anticancer therapy or heart-relevant radiotherapy between 1976–2019. We invited them for an echocardiographic assessment including 2-dimensional LVEF (reduced LVEF: <54% for females; <52% for males). We stratified CCS in anthracyclines only, heart-relevant radiotherapy only, both, and a standard risk group exposed to any other systemic anticancer treatment and compared groups using ANOVA- and t-tests. We performed multivariable linear regression to investigate sociodemographic, treatment-, and lifestyle-related factors (hypertension, smoking, dyslipidemia, diabetes, overweight/obesity), and multilevel modelling to assess LVEF changes over time.

**Results:** We assessed 487 CCS (median age: 32 years, IQR: 24–39) with a median time since diagnosis of 24 years (IQR 17‒31). Overall prevalence of reduced LVEF was 7.2% (35/487), 7.3% (19/260) for anthracyclines only, 3.6% (1/28) for heart-relevant radiotherapy only, 9.4% (8/85) for both, and 6.1% (7/114) for the standard risk group. Male CCS were at risk for reduced LVEF [β coefficient for male sex = −1.37; 95%CI −2.36, −0.37], and higher cumulative anthracycline dose (per 100 mg/m²) was associated with reduced LVEF (β −0.83; 95%CI −1.20, −0.45). Heart-relevant radiotherapy, other systemic anticancer treatments, and lifestyle were not associated with LVEF. We found a slight annual improvement of LVEF 0.24% among 140 participants followed up for a median of 4.3 years, however, there was no indication of statistical significance (p=0.12).

**Conclusion:** This first nationwide study found that a substantial proportion of survivors had reduced LVEF, especially males and those treated with anthracyclines. While cardiac function appeared stable over a four-year period, long-term changes remain uncertain, emphasizing the importance of ongoing cardiac surveillance in this vulnerable population.

## Introduction

Childhood cancer survival has improved significantly over recent decades; it now exceeds 85% in Switzerland^1^. The growing population of childhood cancer survivors (CCS) is at increased risk of long-term morbidity and mortality, with cardiovascular disease representing the leading nonmalignant cause of death^2^. CCS treated with high cumulative doses of anthracyclines or heart-relevant radiotherapy are at the highest risk of developing heart failure^3^. Other cardiovascular risk factors further potentiate their cardiovascular morbidity^4^. For this group of survivors, the International Guideline Harmonization Group (IGHG) recommends regular surveillance of left ventricular ejection fraction (LVEF) by echocardiography^3^. Yet cardiac abnormalities have been reported even in CCS who did not receive anthracyclines or heart-relevant radiation ^5^. In comparison with siblings, a two- to three-fold increased risk of cardiac dysfunction has been observed in survivors without exposure to these known cardiotoxic treatments^6^. These findings suggest that other systemic anticancer treatments such as alkylating agents, vincristine, cisplatin, corticosteroids, and hematopoietic stem cell transplantation (HSCT) also contribute to long-term cardiotoxicity. Understanding the late effects of these treatments is essential for improving individual risk prediction and tailoring surveillance strategies.

Despite established recommendations, the optimal surveillance strategy for cardiomyopathy in CCS remains uncertain. Most research assessing LVEF in CCS has been based on cross-sectional study designs typically assessing participants only once, many years after treatment^7^. While these studies provide evidence on the prevalence of reduced LVEF, they cannot capture the progression of cardiac dysfunction over time. Whether and how cardiac dysfunction evolves after childhood cancer treatment remains unclear. Moreover, current IGHG recommendations are largely based on data from North America and the Netherlands, which may not be applicable to Swiss CCS due to differences in treatment protocols and lifestyle^6,8–10^.

To address these evidence gaps, we conducted the first longitudinal echocardiography study of adult CCS in Switzerland to 1) assess the prevalence of reduced LVEF and its associated factors including known cardiotoxic and other systemic anticancer treatments, and 2) investigate longitudinal changes of LVEF to better characterize its dynamics after childhood cancer treatment.

## Methods

### Study design and participants

The CardioOnco study is an ongoing, multicenter longitudinal cohort study assessing cardiovascular health in adult CCS across Switzerland who are registered in the Swiss Childhood Cancer Registry^11,12^. Since 1976, the ChCR has included all persons diagnosed with cancer in Switzerland before the age of 20, with cases classified according to the International Classification of Childhood Cancer, Third Edition (ICCC-3)^13,14^. The study was launched as a single-center study in 2016 and expanded into a multicenter study, now involving five participating centers. Inclusion criteria are age ≥18 years; treatment in one of the study centers with any chemotherapy, and/or heart-relevant radiotherapy; survival at least five years since diagnosis; registered in the ChCR; and residence in Switzerland^11,12^. The exclusion criterion is treatment with only surgery and/or radiotherapy not involving the heart (these survivors are considered at low risk for treatment-related cardiac dysfunction). Study teams at each center identified eligible CCS and sent postal invitations to attend a cardio-oncology outpatient clinic for baseline assessment that included transthoracic echocardiography (TTE) evaluation, physical examination, and taking medical history. Participants exposed to anthracyclines and/or heart-relevant radiotherapy were scheduled for follow-up visits every 2 to 5 years depending on their cumulative doses^3^. Participants not exposed to anthracyclines or heart-relevant radiotherapy but treated with any other systemic anticancer treatments were followed longitudinally if clinically indicated. We collected written informed consent from all participants. The study was conducted in accordance with the Declaration of Helsinki and received ethical approval from the Ethics Committee of the Canton of Bern (KEK-BE: 2017-01612). Detailed information regarding the study design was published previously^15^. This study has the ClinicalTrials.gov identifier NCT03790943.

## Explanatory variables

### Sociodemographics

We received information about sex, age, and primary treatment institution from the ChCR. During the visit, we asked patients about their living situation, marital status, and profession. We coded those with a current occupation into the European Socioeconomic Classification (ESeC). We categorized occupational position as low (lower clerical, services and sales workers, skilled workers, and semi-skilled and unskilled workers [ESeC class 7, 8, and 9]), intermediate (small employers and self-employed, farmers, lower supervisors, and technicians [ESeC class 4, 5, and 6]), or high (higher professionals and managers, higher clerical, services, and sales workers [ESeC class 1, 2, and 3])^16^.

### Cardiovascular risk factors

We asked patients during the visit about cardiovascular risk factors including diabetes mellitus, hypertension, dyslipidemia, and smoking status. We measured blood pressure three times in a sitting position with a break of one minute in between measurements and took the average of the last two readings ^17^. We defined hypertension if the mean systolic blood pressure was ≥140 mmHg, the diastolic blood pressure was ≥90 mmHg, or if the patient took prescribed antihypertensive medication^17^. We further measured height and weight to obtain survivor body mass index (BMI), and measured waist-hip-ratio^15^. BMI was calculated as weight in kilograms divided by height in meters squared and classified according to WHO criteria: underweight (<18.5 kg/m²), normal weight (18.5–24.9 kg/m²), overweight (25–29.9 kg/m²), and obese (≥30 kg/m²)^18^. Waist-hip ratio was categorized as nonobese (male <0.9, female <0.85) or obese (male ≥0.9, female ≥0.85) based on established sex-specific thresholds^19^.

### Cancer history and treatment

The ChCR provided clinical information about cancer diagnosis, date of diagnosis, and relapse history. We collected data from the ChCr and from medical records on cancer treatments and doses of anthracyclines, alkylating agents, cisplatin, vincristine, steroids, heart-relevant radiotherapy, and conditioning regimens of HSCT. To quantify anthracycline exposure, we converted doses of daunorubicin, idarubicin, epirubicin, and mitoxantrone into doxorubicin isotoxic equivalents following the latest IGHG recommendations^20,21^. We defined heart-relevant radiotherapy as any radiotherapy targeting the chest, abdomen, thoracic or whole spine, and collected the highest reported dose in Grays (Gy) to any of these fields. If total body irradiation (TBI) was additionally administered, we added the respective dose to the highest field dose. We calculated cyclophosphamide equivalent doses based on the total amounts of cyclophosphamide and ifosfamide following COG recommendations^21^. We determined prednisone equivalent doses based on the total amount of prednisone and dexamethasone using a previously published conversion method^22^.

Risk groups were defined based on exposure to anthracyclines (including mitoxantrone), heart-relevant radiotherapy, both anthracyclines and heart-relevant radiotherapy, and a standard risk group exposed to any other systemic anticancer treatment including alkylating agents, vincristine, cisplatin, corticosteroids, and HSCT.

### Outcome measures

All echocardiograms were conducted by experienced cardiologists affiliated with the cardiology departments of the participating centers using GE Vivid E9, E90 or E95 (GE Vingmed Ultrasound, Horten, Norway) and Philips EPIQ 7 ultrasound scanners (Philips Ultrasound, Inc., Bothell, Washington, USA). All cardiologists followed standardized imaging protocols to ensure consistency across sites and were blinded with respect to participants cancer treatment whenever possible^23^. To assess LVEF, we used the two-dimensional Simpson’s biplane method^23,24^. We defined reduced LVEF with sex-specific cut-off values following international recommendations (men < 52%; women: <54%)^23^. For patients with an LVEF below 50%, we reviewed medical records and extracted information on potential interventions aimed at improving LVEF including prescribed medications and lifestyle modifications.

### Statistical analysis

We summarized continuous variables using the mean (standard deviation) or median (interquartile range), and categorical variables were presented as number (percentages). We reported the prevalence of reduced LVEF for all risk groups. We compared groups using t-tests and analysis of variance (ANOVA), and used univariable and multivariable linear regression models to identify associations between explanatory variables and reduced LVEF. In the multivariable models, we used the a priori factors sex, age, and cumulative doses of radiotherapy and anthracyclines, which have been associated with cardiac dysfunction in CCS^3^. We also chose additional factors that include cancer diagnosis, treatment with cyclophosphamide, vincristine, cisplatin, steroids, and HSCT, and clinical risk factors such as diabetes, dyslipidemia, hypertension, smoking, and waist-hip ratio. These additional factors were included in the multivariable model if they had a p-value < 0.2 in univariable analysis. We also conducted multilevel linear regression modeling to assess changes in LVEF over time (details of the model are provided in Supplementary Materials). To assess potential selection bias, we compared characteristics of our study participants with those of nonparticipants, which in both cases we obtained from the ChCR. This modeling approach allowed us to capture both the average change of LVEF over time and subject-specific deviations from this change. We performed all analyses using R Statistical Software (V-4.5.0.; R Core Team 2024).

## Results

### Characteristics of Study Population

Between November 2017 and December 2024, we invited 1297 eligible CCS and measured LVEF in 487 (38%) (Supp Figure 1). Median age at study was 32 years (IQR 24–39 years) with a median time since diagnosis of 24 years (IQR 17–31 years) (Tables I and II). The most frequent cancer diagnoses were leukemias (44%), followed by lymphomas (21%), malignant bone tumors (7%), and renal tumors (7%). Among all participants, 64% had received anthracyclines with a median cumulative dose of 154mg/m2 (standard deviation [SD] 145mg/m2), 23% had received heart-relevant radiotherapy with a median cumulative dose of 26 Gy (SD 13Gy), 7% received autologous or allogeneic hematopoietic stem cell transplantation (HSCT) and 23% had none of these.

**Figure 1:**
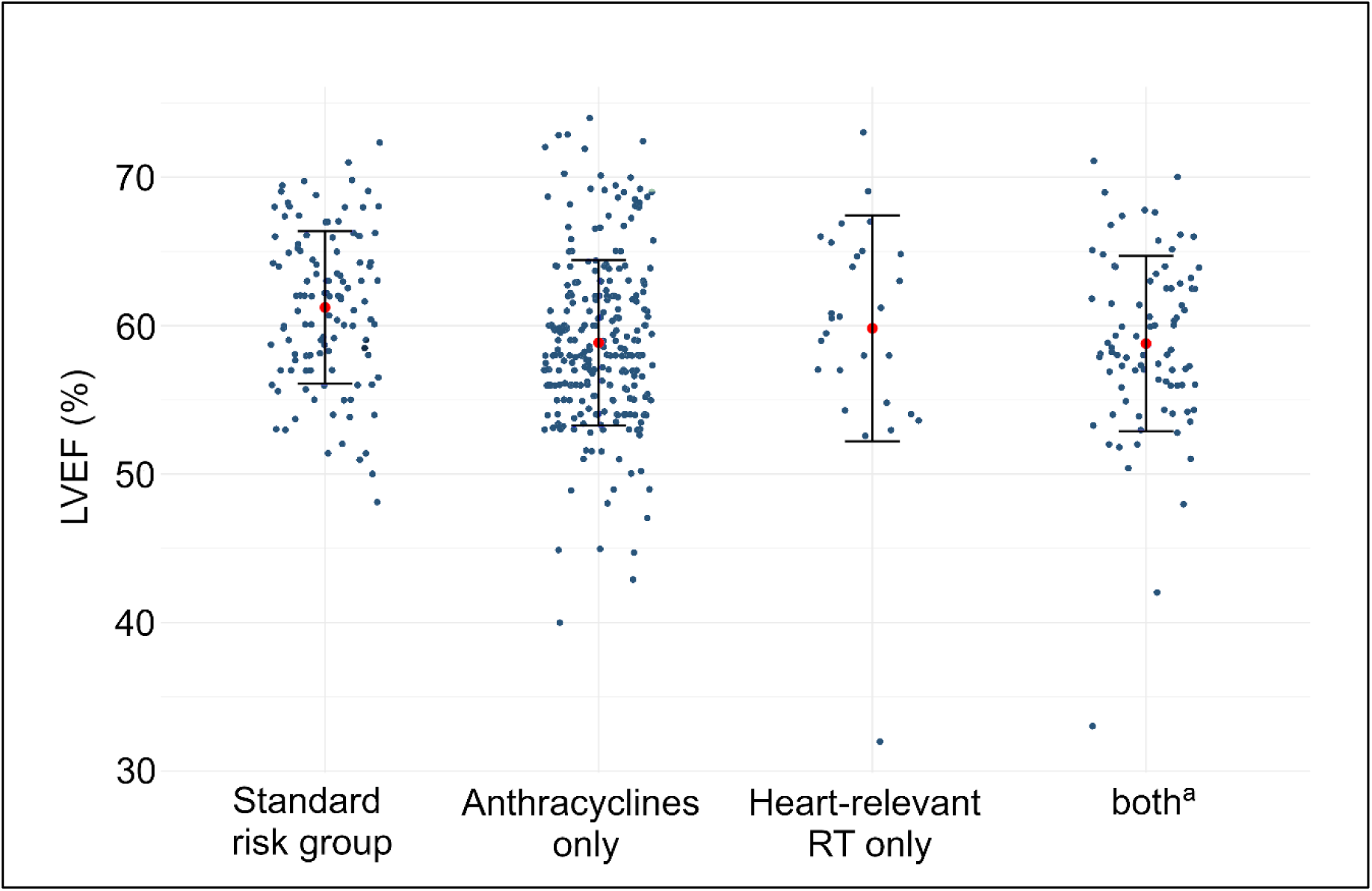
Distribution of left ventricular ejection fraction among CardioOnco Study participants stratified by risk group ***Note:*** *Each point represents an individual childhood cancer survivor (CSS). Red points indicate the group mean LVEF, with error bars representing the 95% confidence intervals*. *Abbreviations: LVEF, left ventricular ejection fraction; AC, anthracyclines; RT, radiotherapy; CCS, childhood cancer survivor* ^a^Survivors exposed to anthracyclines and heart-relevant radiotherapy.

**Table I:**
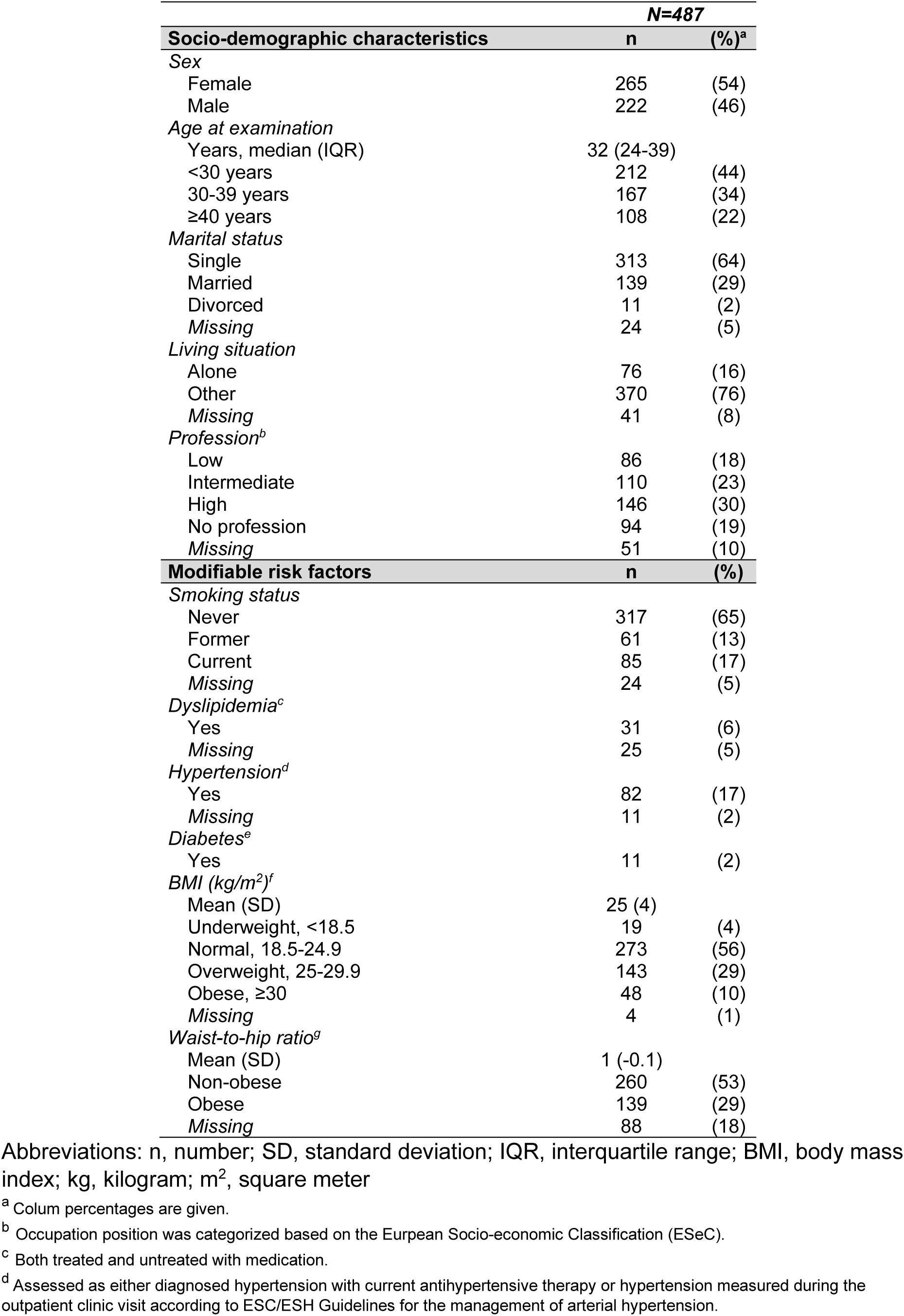
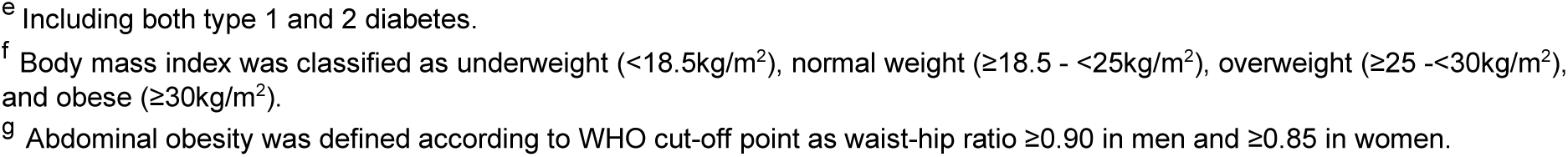
Socio-demographic and cardiovascular risk factors of CardioOnco study participants.

| *N=487* |  |  |
| --- | --- | --- |
| **Socio-demographic characteristics** | **n** | **(%)<sup>a</sup>** |
| *Sex* |  |  |
| Female | 265 | (54) |
| Male | 222 | (46) |
| *Age at examination* |  |  |
| Years, median (IQR) | 32 (24-39) |  |
| <30 years | 212 | (44) |
| 30-39 years | 167 | (34) |
| ≥40 years | 108 | (22) |
| *Marital status* |  |  |
| Single | 313 | (64) |
| Married | 139 | (29) |
| Divorced | 11 | (2) |
| Missing | 24 | (5) |
| *Living situation* |  |  |
| Alone | 76 | (16) |
| Other | 370 | (76) |
| Missing | 41 | (8) |
| *Profession<sup>b</sup>* |  |  |
| Low | 86 | (18) |
| Intermediate | 110 | (23) |
| High | 146 | (30) |
| No profession | 94 | (19) |
| Missing | 51 | (10) |
| **Modifiable risk factors** | **n** | **(%)** |
| *Smoking status* |  |  |
| Never | 317 | (65) |
| Former | 61 | (13) |
| Current | 85 | (17) |
| Missing | 24 | (5) |
| *Dyslipidemia<sup>c</sup>* |  |  |
| Yes | 31 | (6) |
| Missing | 25 | (5) |
| *Hypertension<sup>d</sup>* |  |  |
| Yes | 82 | (17) |
| Missing | 11 | (2) |
| *Diabetes<sup>e</sup>* |  |  |
| Yes | 11 | (2) |
| *BMI (kg/m<sup>2</sup>)<sup>f</sup>* |  |  |
| Mean (SD) | 25 (4) |  |
| Underweight, <18.5 | 19 | (4) |
| Normal, 18.5-24.9 | 273 | (56) |
| Overweight, 25-29.9 | 143 | (29) |
| Obese, ≥30 | 48 | (10) |
| Missing | 4 | (1) |
| *Waist-to-hip ratio<sup>g</sup>* |  |  |
| Mean (SD) | 1 (-0.1) |  |
| Non-obese | 260 | (53) |
| Obese | 139 | (29) |
| Missing | 88 | (18) |

Abbreviations: n, number; SD, standard deviation; IQR, interquartile range; BMI, body mass index; kg, kilogram; m<sup>2</sup>, square meter
<sup>a</sup> Column percentages are given.
<sup>b</sup> Occupation position was categorized based on the European Socio-economic Classification (ESeC).
<sup>c</sup> Both treated and untreated with medication.
<sup>d</sup> Assessed as either diagnosed hypertension with current antihypertensive therapy or hypertension measured during the outpatient clinic visit according to ESC/ESH Guidelines for the management of arterial hypertension.
<sup>e</sup> Including both type 1 and 2 diabetes. <sup>f</sup> Body mass index was classified as underweight (<18.5kg/m<sup>2</sup>), normal weight (≥18.5 - <25kg/m<sup>2</sup>), overweight (≥25 -<30kg/m<sup>2</sup>), and obese (≥30kg/m<sup>2</sup>). <sup>g</sup> Abdominal obesity was defined according to WHO cut-off point as waist-hip ratio ≥0.90 in men and ≥0.85 in women.

**Table II:**
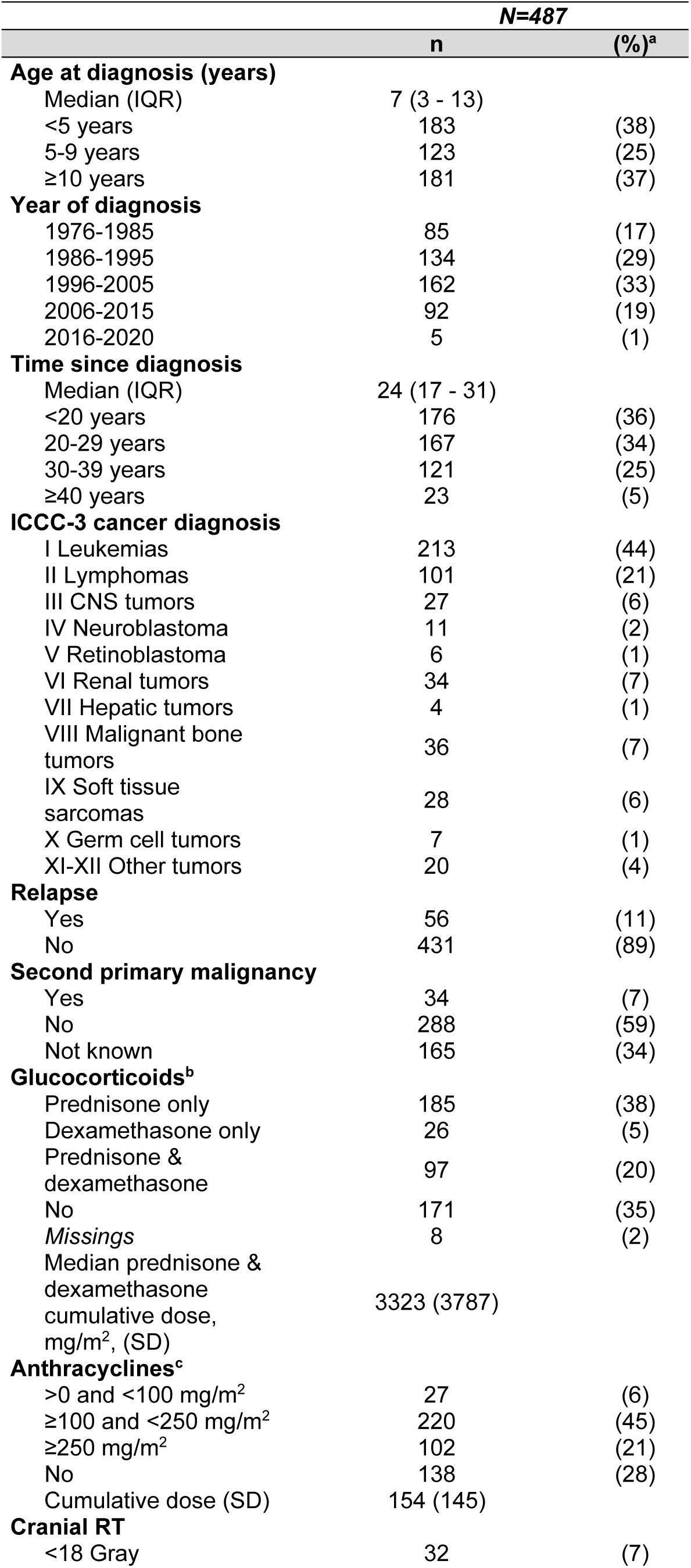
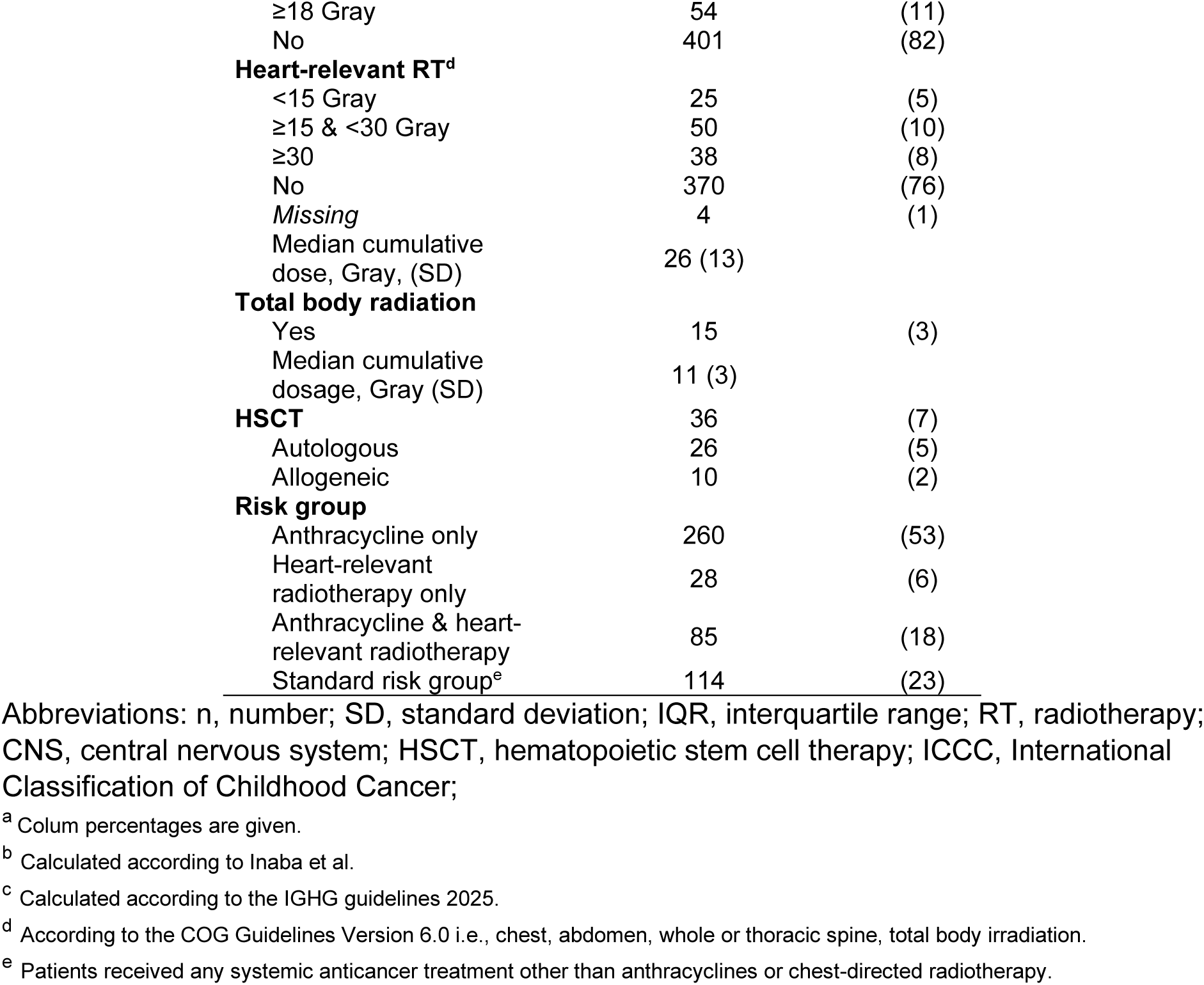
Cancer history and treatment of CardioOnco study participants.

| **N=487** |  |  |
| --- | --- | --- |
|  | **n** | **(%)<sup>a</sup>** |
| **Age at diagnosis (years)** |  |  |
| Median (IQR) | 7 (3 - 13) |  |
| <5 years | 183 | (38) |
| 5-9 years | 123 | (25) |
| ≥10 years | 181 | (37) |
| **Year of diagnosis** |  |  |
| 1976-1985 | 85 | (17) |
| 1986-1995 | 134 | (29) |
| 1996-2005 | 162 | (33) |
| 2006-2015 | 92 | (19) |
| 2016-2020 | 5 | (1) |
| **Time since diagnosis** |  |  |
| Median (IQR) | 24 (17 - 31) |  |
| <20 years | 176 | (36) |
| 20-29 years | 167 | (34) |
| 30-39 years | 121 | (25) |
| ≥40 years | 23 | (5) |
| **ICCC-3 cancer diagnosis** |  |  |
| I Leukemias | 213 | (44) |
| II Lymphomas | 101 | (21) |
| III CNS tumors | 27 | (6) |
| IV Neuroblastoma | 11 | (2) |
| V Retinoblastoma | 6 | (1) |
| VI Renal tumors | 34 | (7) |
| VII Hepatic tumors | 4 | (1) |
| VIII Malignant bone tumors | 36 | (7) |
| IX Soft tissue sarcomas | 28 | (6) |
| X Germ cell tumors | 7 | (1) |
| XI-XII Other tumors | 20 | (4) |
| **Relapse** |  |  |
| Yes | 56 | (11) |
| No | 431 | (89) |
| **Second primary malignancy** |  |  |
| Yes | 34 | (7) |
| No | 288 | (59) |
| Not known | 165 | (34) |
| **Glucocorticoids<sup>b</sup>** |  |  |
| Prednisone only | 185 | (38) |
| Dexamethasone only | 26 | (5) |
| Prednisone & dexamethasone | 97 | (20) |
| No | 171 | (35) |
| *Missings* | 8 | (2) |
| Median prednisone & dexamethasone cumulative dose, mg/m<sup>2</sup>, (SD) | 3323 (3787) |  |
| **Anthracyclines<sup>c</sup>** |  |  |
| >0 and <100 mg/m<sup>2</sup> | 27 | (6) |
| ≥100 and <250 mg/m<sup>2</sup> | 220 | (45) |
| ≥250 mg/m<sup>2</sup> | 102 | (21) |
| No | 138 | (28) |
| Cumulative dose (SD) | 154 (145) |  |
| **Cranial RT** |  |  |
| <18 Gray | 32 | (7) |
| ≥18 Gray | 54 | (11) |
| No | 401 | (82) |
| **Heart-relevant RT<sup>d</sup>** |  |  |
| <15 Gray | 25 | (5) |
| ≥15 & <30 Gray | 50 | (10) |
| ≥30 | 38 | (8) |
| No | 370 | (76) |
| *Missing* | 4 | (1) |
| Median cumulative dose, Gray, (SD) | 26 (13) |  |
| **Total body radiation** |  |  |
| Yes | 15 | (3) |
| Median cumulative dosage, Gray (SD) | 11 (3) |  |
| **HSCT** | 36 | (7) |
| Autologous | 26 | (5) |
| Allogeneic | 10 | (2) |
| **Risk group** |  |  |
| Anthracycline only | 260 | (53) |
| Heart-relevant radiotherapy only | 28 | (6) |
| Anthracycline & heart-relevant radiotherapy | 85 | (18) |
| Standard risk group<sup>e</sup> | 114 | (23) |

Abbreviations: n, number; SD, standard deviation; IQR, interquartile range; RT, radiotherapy; CNS, central nervous system; HSCT, hematopoietic stem cell therapy; ICC, International Classification of Childhood Cancer;
<sup>a</sup> Column percentages are given.
<sup>b</sup> Calculated according to Inaba et al.
<sup>c</sup> Calculated according to the ICHG guidelines 2025.
<sup>d</sup> According to the COG Guidelines Version 6.0 i.e., chest, abdomen, whole or thoracic spine, total body irradiation.
<sup>e</sup> Patients received any systemic anticancer treatment other than anthracyclines or chest-directed radiotherapy.

Cardiovascular risk factors were common: 30% were former or current smokers, 17% had hypertension, and 6% had dyslipidemia. A diagnosis of type 1 or type 2 diabetes mellitus was reported in 2% of participants. Median BMI was 23.9 kg/m², and 39% of participants were classified as overweight or obese. To assess potential selection bias, we compared study participants with nonparticipants (those invited but who did not participate). Both groups were largely comparable in terms of age, sex, diagnosis, and treatment characteristics, with statistically significant differences observed only in the proportion of survivors who received chemotherapy and in the distribution of germ cell and other tumors (Supplementary Table I).

### Echocardiographic characteristics

The mean LVEF across all participants was 59.5% (SD 5.7%) (Figure 1 & Supp. Table II). When stratified by risk groups, patients treated with anthracycline only exhibited the lowest mean LVEF (58.8±5.6%), while the highest mean LVEF was observed among survivors in the standard risk group (61.2±5.1%). When we compared individual risk groups against the standard risk group, the survivors treated with anthracyclines only (58.8±5.6%, p<0.01) and those treated with a combination of anthracyclines and heart-relevant radiotherapy (58.8±5.9%, p<0.01) had lower mean LVEF.

### Prevalence of reduced LVEF

Among all participants, 7.2% had a reduced LVEF (Supp Table II). Reduced LVEF was most common in survivors treated with both anthracyclines and heart-relevant radiotherapy (9.4%), and least common in survivors treated with heart-relevant radiotherapy alone (3.6%) (Figure 2). Among survivors in the standard risk group, most reduced LVEF values were clustered near the lower limit of normal. The prevalence of reduced LVEF did not differ significantly between risk groups (ANOVA, p=0.68).

**Figure 2:**
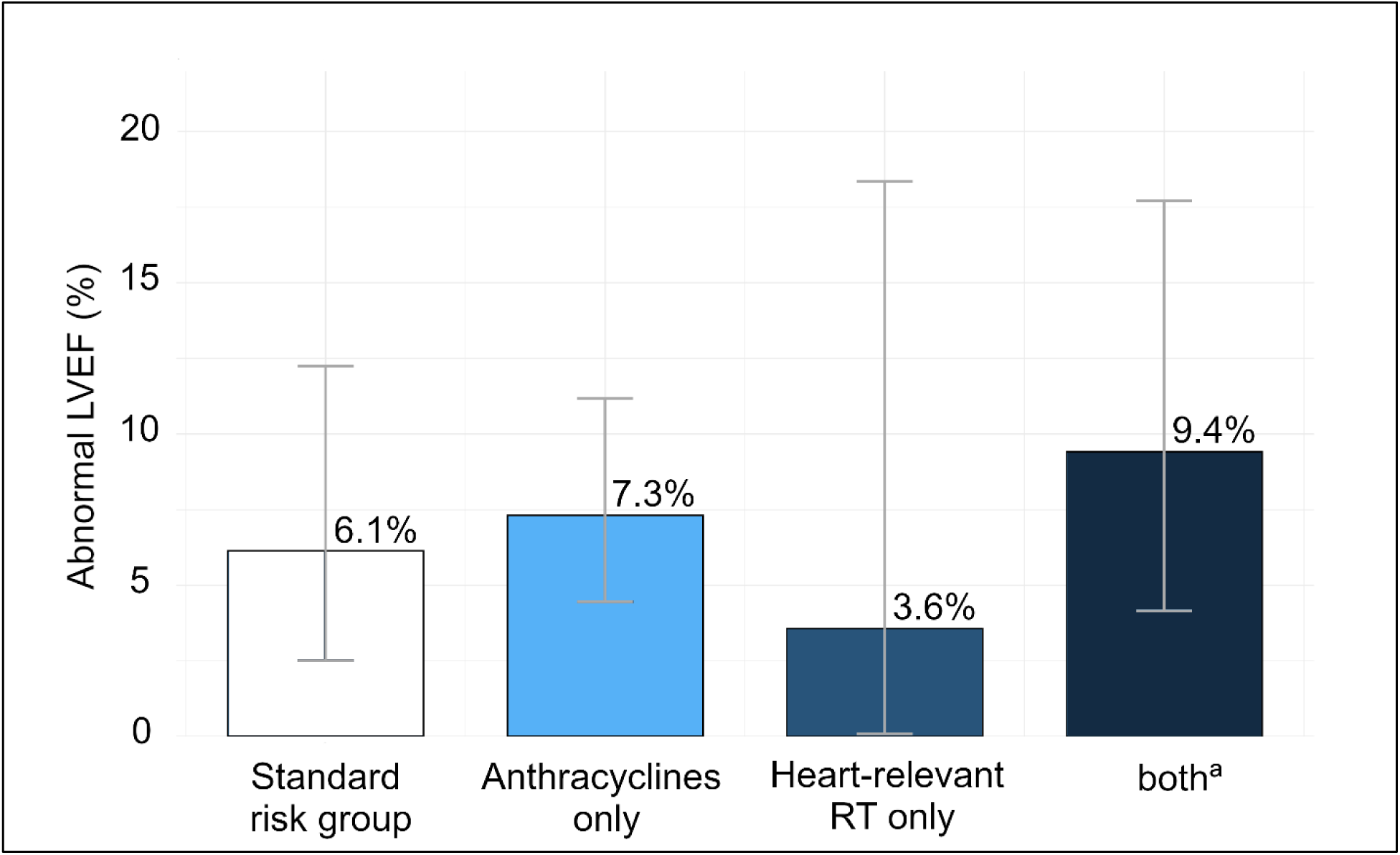
Prevalence of abnormal left ventricular ejection fraction among CardioOnco study participants stratified by risk group ***Note****: Bars represent the proportion of survivors with abnormal LVEF in each group; error bars indicate 95% confidence intervals*. *Abbreviations: LVEF, left ventricular ejection fraction; AC anthracyclines; RT radiotherapy; CCS, childhood cancer survivors* ^a^Survivors exposed to anthracyclines and heart-relevant radiotherapy.

### Factors associated with LVEF

Survivors treated with anthracyclines and male survivors were at risk for lower LVEF (unstandardized coefficient (β) for cumulative dose of anthracyclines = −0.83; 95% CI −1.20.– −0.45; per 100mg/m^2^; β for male sex= −1.37; 95%CI −2.36, −0.37) (Table III). Survivors treated with heart-relevant radiotherapy, other systemic anticancer treatments, and the presence of cardiovascular risk factors were not associated with LVEF.

**Table III:**
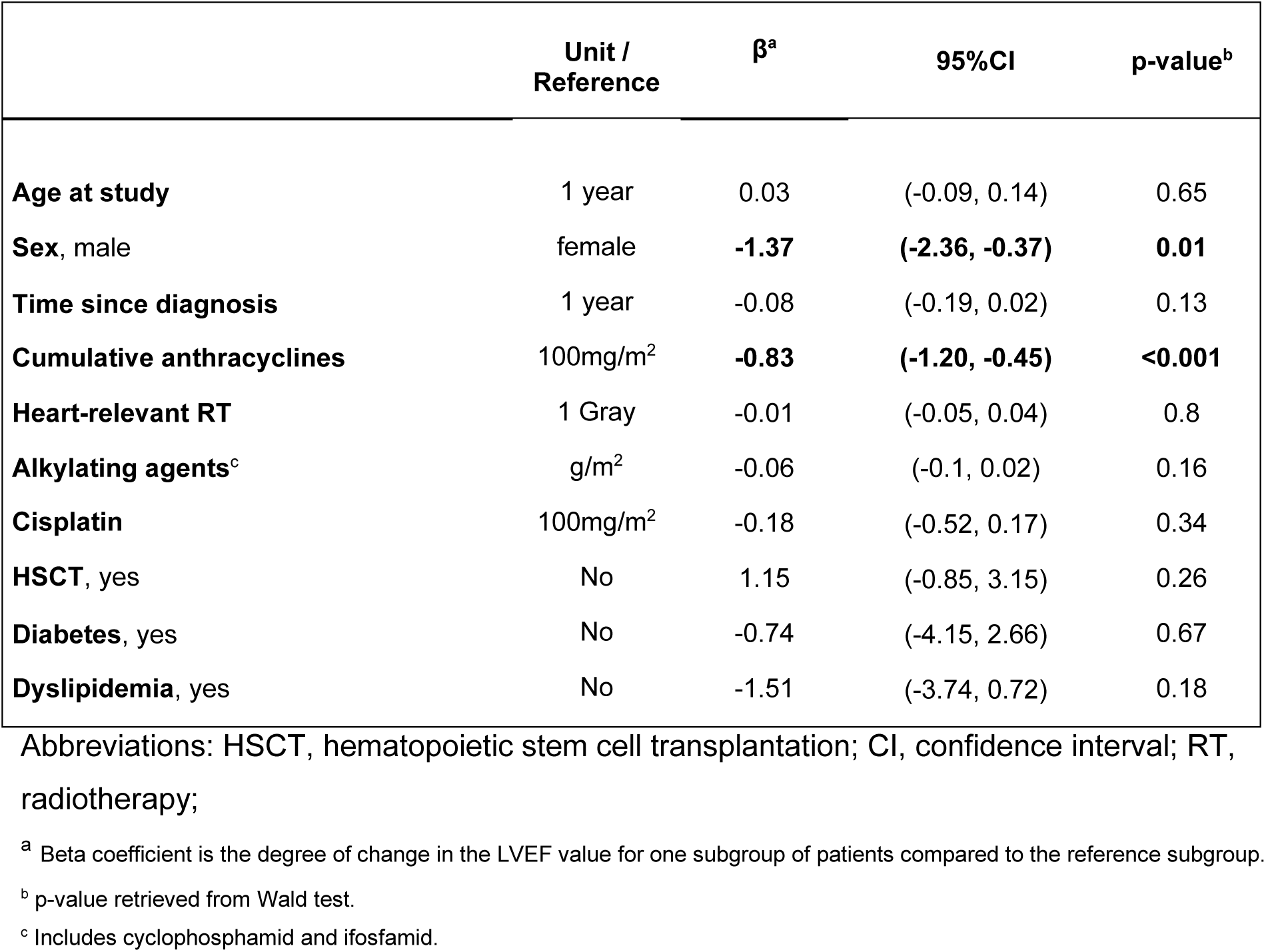
Factors associated with left ventricular ejection fraction among CardioOnco participants using multivariable linear regression (n=480)

| | Unit /<br>Reference | $\beta^a$ | 95%CI | p-value<sup>b</sup> |
| --- | --- | --- | --- | --- |
| **Age at study** | 1 year | 0.03 | (-0.09, 0.14) | 0.65 |
| **Sex, male** | female | **-1.37** | **(-2.36, -0.37)** | **0.01** |
| **Time since diagnosis** | 1 year | -0.08 | (-0.19, 0.02) | 0.13 |
| **Cumulative anthracyclines** | 100mg/m<sup>2</sup> | **-0.83** | **(-1.20, -0.45)** | **<0.001** |
| **Heart-relevant RT** | 1 Gray | -0.01 | (-0.05, 0.04) | 0.8 |
| **Alkylating agents<sup>c</sup>** | g/m<sup>2</sup> | -0.06 | (-0.1, 0.02) | 0.16 |
| **Cisplatin** | 100mg/m<sup>2</sup> | -0.18 | (-0.52, 0.17) | 0.34 |
| **HSCT, yes** | No | 1.15 | (-0.85, 3.15) | 0.26 |
| **Diabetes, yes** | No | -0.74 | (-4.15, 2.66) | 0.67 |
| **Dyslipidemia, yes** | No | -1.51 | (-3.74, 0.72) | 0.18 |

Abbreviations: HSCT, hematopoietic stem cell transplantation; CI, confidence interval; RT, radiotherapy;
<sup>a</sup> Beta coefficient is the degree of change in the LVEF value for one subgroup of patients compared to the reference subgroup.
<sup>b</sup> p-value retrieved from Wald test.
<sup>c</sup> Includes cyclophosphamid and ifosfamid.

### Longitudinal assessment of LVEF using multilevel modelling

Among 140 patients followed longitudinally, 133 (95%) received anthracyclines, heart-relevant radiotherapy or both (Supp. Table III). Overall, we observed an average annual increase of LVEF 0.24%, with no indication of statistical significance (p=0.12) (Figure 3). We found no evidence of a meaningful change in LVEF over time and no significant differences between individual patients were detected.

**Figure 3:**
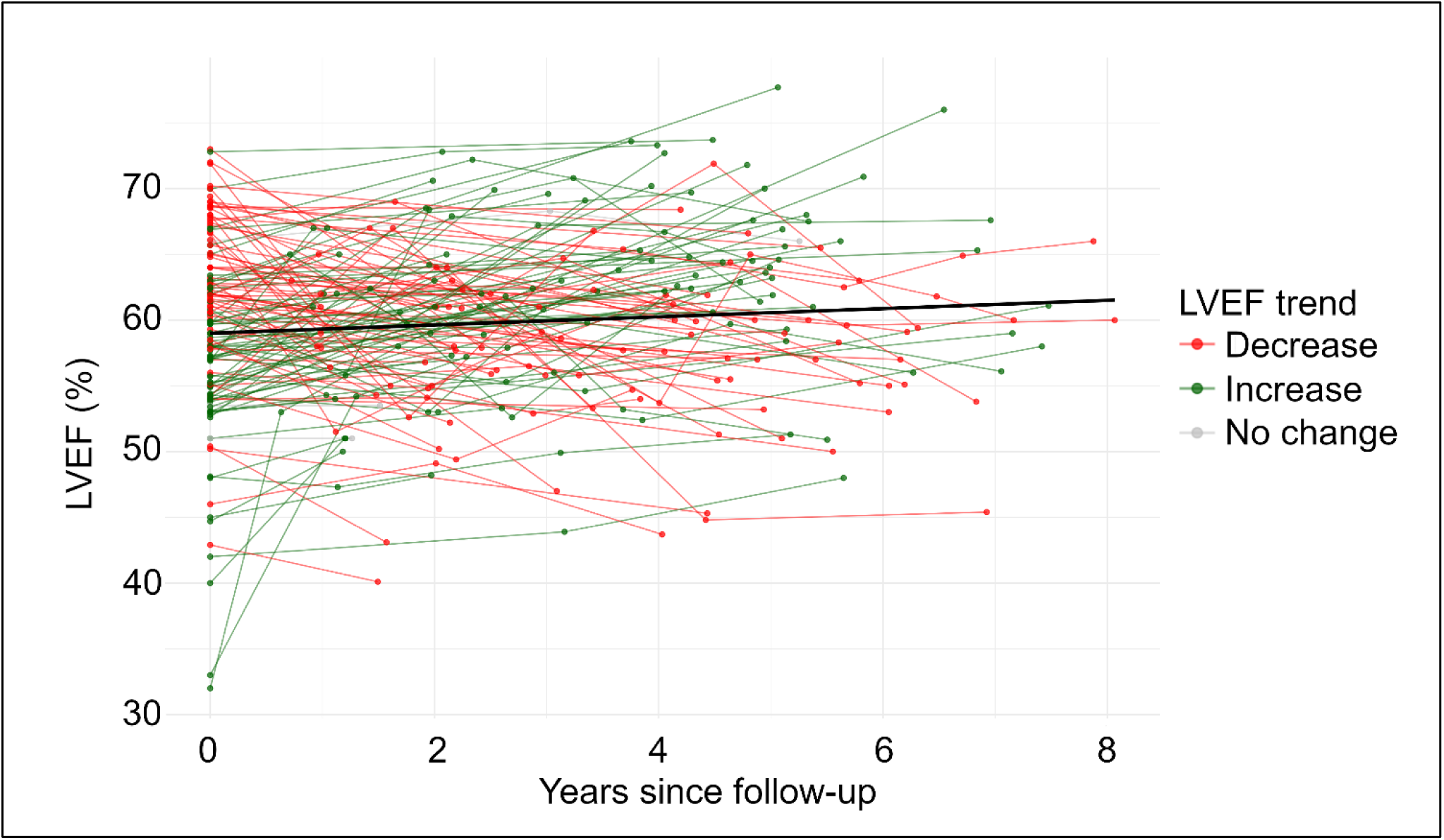
Longitudinal assessment of left ventricular ejection fraction among CardioOnco study participants (n=140) **Note**: In total, 140 participants were assessed longitudinally with a median follow-up time of 4.3 years. The mean LVEF at baseline assessment was 59.2% and improved by 0.24% LVEF per year, although not statistically significant (p=0.12). *Abbreviations: LVEF, left ventricular ejection fraction; n, number;*

Among 15 patients with LVEF below 50%, we observed potential interventions that may have improved LVEF during follow-up. Cardiologists started heart failure medication in CCS with LVEF <40%, addressed modifiable cardiovascular risk factors such as hypertension, diabetes, and dyslipidemia, and provided lifestyle counselling including engagement in physical activity, weight reduction, and smoking cessation when appropriate.

## Discussion

### Summary of findings

This nationwide, longitudinal cohort study assessing cardiac function among adult CCS in Switzerland found that 7.2% of survivors showed signs of cardiac dysfunction assessed as reduced LVEF, especially male survivors and those treated with higher cumulative anthracycline doses. We found no evidence of a change in LVEF over time.

### Prevalence of reduced LVEF

In population-based cohorts of healthy adults, reduced LVEF has been reported in 1.7–3.6%, according to LVEF cut-offs of <50% and <55% respectively ^25^. These prevalences are derived from older populations (mean age ∼ 61 years), underscoring the importance of the 7.2% observed in our young CCS cohort^25^. This prevalence is comparable to the 5.8% reported in the largest CCS study to date, the St. Jude Lifetime Cohort (n = 1820)^8^. However, reported prevalences in the literature vary substantially, ranging from 0.8% to 24.2% across seven studies published since 2012^6,8,9,26–29^. These differences may in part reflect different intervals between cancer diagnosis and outcome assessment. The study reporting the lowest prevalence (0.8%) had the shortest follow-up period (9 years), while the highest prevalence (24.2%) was reported by the study with the longest time since diagnosis (27 years)^6,9^. These observations suggest that cardiotoxic effects may become more apparent with longer follow-up, which emphasizes the importance of extended cardiac surveillance in this population. Another factor contributing to the variability is the use of different cut-off values for defining reduced LVEF. Although sex-specific thresholds (<54% for females, <52% for males) were introduced by ASE/EACVI in 2016, not all studies have adopted them, with earlier research often using general cut-offs between 50% and 55%^8,9,26–30^. To improve comparability, future studies should consistently apply these standardized criteria.

### Factors associated with reduced LVEF

CCS treated with higher cumulative anthracycline dose had higher risk for reduced LVEF, which is consistent with evidence from the St. Jude Lifetime Cohort study and the Dutch LATER study, the only two studies that used regression analysis to assess factors associated with LVEF^6,8^. Anthracyclines induce myocyte apoptosis through complex formation with topoisomerase-IIβ and DNA, providing a mechanistic explanation for their cardiotoxic effect^31^. The IGHG and European Society of Cardiology guidelines acknowledge the crucial role of anthracyclines in cardiac dysfunction and recommend more frequent surveillance for CCS exposed to high cumulative doses^3,32^. Although the IGHG also recommends screening for CCS exposed to heart-relevant radiotherapy, we found no association with reduced LVEF. Both the St. Jude and Dutch studies reported stronger associations between survivors treated with chest-directed radiotherapy and abnormal global longitudinal strain (GLS) than with LVEF^6,8^, suggesting GLS may be more sensitive in detecting radiotherapy-induced cardiotoxicity. Another explanation for our findings may lie in how we defined radiotherapy exposure. We used the highest reported dose to heart-relevant fields, whereas the St. Jude and Dutch study applied organ-specific dose reconstruction – a method that accounts for radiation orientation, field size, and patient anatomy, which is crucial since the proximity of radiation field to relevant organs can vary substantially with body size^6,8,21,33^. Our approach likely introduced uncertainty in cardiac dose estimation, possibly contributing to the low prevalence of reduced LVEF (3.6%) in the radiotherapy group. Furthermore, we found no association between CCS exposed to other systemic anticancer treatments and LVEF. Evidence regarding cardiac effects of these treatments is conflicting. Subtle cardiac changes that included increased left ventricular mass and wall thickness were observed in 201 CCS survivors not exposed to treatments known to be cardiotoxic compared to siblings⁵. The clinical relevance of these cardiac changes remains questionable because most studies have not shown an association between CCS exposed to these treatments and heart failure^34,35^. Further research is needed to determine whether these subtle changes represent silent residuals or may progress to clinically manifest disease.

We identified male CCS as being at higher risk for reduced LVEF. This is surprising since female CCS have been linked to heart failure^34^. The St. Jude study also found increased risk in males, while the Dutch study reported female sex as a risk factor^6,8^. This inconsistency may reflect differences in LVEF definitions: both studies conducted nonlinear regression models with reduced LVEF defined as <50% by the St. Jude study and a sex-specific cut-off by the Dutch study (<52% for males, <54% for females)^6,8^. The sex-specific cut-off is recommended by the ASE/EACVI and reflects normal physiological differences, with females having slightly higher LVEF values than males^30^. Since we used LVEF as a continuous outcome in a linear model, our finding may reflect these physiological sex differences and should be interpreted with caution.

We did not identify an association between survivors with modifiable cardiovascular risk factors and reduced LVEF. Similarly, both the St. Jude and Dutch study found no association with reduced LVEF, with the exception of hypertension, which was associated with lower LVEF in the St. Jude study^6,8^. However, both studies reported stronger and more consistent associations between modifiable risk factors and abnormal GLS. Because the median age in these cohorts is ∼30 years, the impact of modifiable cardiovascular risk factors may emerge later in life, and GLS, as a more sensitive marker, may already detect subtle dysfunction not yet evident in LVEF. Further studies with longer follow-up are needed to clarify the long-term impact of modifiable risk factors.

### Longitudinal assessment

We observed no changes in LVEF over time. Although there was a slight annual increase in LVEF, it was not statistically significant. While regression to the mean cannot be entirely excluded, it is unlikely to have influenced the findings on this study. Cardiotoxicity is known to progress slowly, and the median follow-up duration of 4.3 years in our cohort may have been too short to detect a decline in LVEF. Interventions prescribed by cardiologists are more likely to show effects within this timeframe and may have contributed to the observed increase in LVEF in some patients during longitudinal follow-up^36^. Longitudinal data on LVEF after childhood cancer remain limited since most studies are cross-sectional. Studies with repeated measurements often focus on prediction modeling rather than assessing changes over time^37,38^. One exception is a US study, which examined longitudinal cardiac changes in a young cohort with a median age of 4.8 years at treatment initiation, which was followed for a median of 11.8 years^39^. The study did not assess LVEF but focused on LV dimensions and contractility, reporting progressive cardiac abnormalities over time^39^. Whether comparable clinical interventions were implemented is unclear and comparability with our findings is limited. Longer follow-up is needed to better understand LVEF changes over time, and the impact of cardiac surveillance and interventions.

### Strengths & limitations

Our nationwide, population-based cohort represents a broad sample of CCS in Switzerland, which overcomes limitations of previous single-center studies and those restricted to specific cancer types or treatments^9,28,29^. We used medical data from the ChCR, allowing for detailed and accurate characterization of cancer diagnoses and treatments. The longitudinal design provides insight into temporal changes in LVEF, an area with limited existing evidence. Nevertheless, as with any long-term follow-up study, survival bias may have distorted our results. CCS who experienced severe early cardiotoxicity leading to cardiac death before study inclusion are not represented in our cohort. However, since heart failure in CCS increases linearly over time, it is unlikely that we missed a substantial number of early cases. Selection bias may also be present, although comparison of participants and nonparticipants using ChCR data showed them to be largely comparable.

While most participants (133/140) were followed in accordance with IGHG recommendations, a subset of individuals from the standard risk group (7/140) were monitored based on clinical indication. These individuals may represent a less healthy subgroup, introducing potential selection bias. Furthermore, the lack of a healthy control group limits comparisons to the general population. Due to the observational design, causal inferences between identified factors and reduced LVEF cannot be made. Finally, echocardiographic images were not independently reviewed. Nonetheless, all measurements were obtained using standardized protocols and comparable ultrasound equipment ensure consistency in image acquisition and interpretation.

### Conclusion

At a median of 32 years after the end of treatment a substantial proportion of CCS who received known cardiotoxic treatments had reduced cardiac function. Male survivors and those exposed to higher cumulative anthracycline doses were at higher risk for reduced LVEF, while neither other systemic anticancer therapies nor modifiable cardiovascular risk factors were associated. Cardiac function remained stable over a four-year time period but longer follow-up is needed to determine long-term trajectories to inform optimal surveillance strategies.

## Supporting information

Supplementary items

## Acknowledgement

We thank all survivors for participating in our study, the study team of the Childhood Cancer Research Group and the team of the Swiss Childhood Cancer Registry.

## Data availability statement

The datasets analyzed during the current study are available from the corresponding author on reasonable request.

## Ethics approval statement

Ethical approval was granted by the Ethics Committee of Canton Bern (KEK-BE: 2017-01612).

