## Supplementary items for "Left ventricular function and its longitudinal change in Swiss childhood cancer survivors – Results from the CardioOnco Study"

**Supp Table I:** Comparison of participants and non-participants of the CardioOnco study

| | Overall (n=1503) | Non-participants (n= 943) | Participants (n= 560) | p-value |
|---|---|---|---|---|
| **Age at invite** | | | | |
| Median (IQR) | 31 (25-38) | 32 (25-38) | 31 (24-38) | 0.14[a] |
| **Sex** | | | | |
| Female | 677 (45%) | 413 (44%) | 264 (47%) | 0.23[b] |
| Male | 826 (55%) | 530 (56%) | 296 (53%) | |
| **Age at diagnosis** | | | | |
| Median (IQR) | 8 (3-13) | 8 (3-13) | 7 (3-13) | 0.31[a] |
| **ICCC-3 cancer diagnosis** | | | | |
| I Leukemias | 592 (40%) | 361 (38%) | 233 (42%) | |
| II Lymphomas | 333 (22%) | 214 (23%) | 115 (21%) | |
| III CNS tumors | 102 (7%) | 74 (8%) | 31 (5%) | |
| IV Neuroblastoma | 47 (3%) | 31 (3%) | 16 (3%) | |
| V Retinoblastoma | 24 (2%) | 18 (2%) | 6 (1%) | |
| VI Renal tumors | 100 (7%) | 55 (6%) | 44 (8%) | **0.01[b]** |
| VII Hepatic tumors | 9 (1%) | 4 (<1%) | 6 (1%) | |
| VIII Malignant bone tumors | 99 (7%) | 53 (6%) | 46 (8%) | |
| IX Soft tissue sarcomas | 101 (7%) | 66 (7%) | 34 (6%) | |
| X Germ cell tumors | 43 (3%) | **32 (3%)[c]** | **9 (2%)[c]** | |
| XI-XII Other tumors | 51 (3%) | **10 (1%)[c]** | **20 (4%)[c]** | |
| **Second primary malignancy** | | | | |
| Yes | 46 (3%) | 26 (3%) | 20 (4%) | 0.46[b] |
| **Chemotherapy** | | | | |
| Yes | 1440 (96%) | **889 (94%)[c]** | **551 (98%)[c]** | **<0.01[b]** |
| **Radiotherapy** | | | | |
| Yes | 541 (36%) | 328 (34.8%) | 213 (38.0%) | 0.22[b] |
| **HSCT** | | | | |
| Yes | 114 (8%) | 79 (8%) | 40 (7%) | 0.42[b] |
| **Relapse** | | | | |
| Yes | 184 (12%) | 113 (12%) | 71 (13%) | 0.75[b] |

Abbreviations: N, number; IQR, interquartile range; HSCT, hematopoietic stem cell transplantation

[a] p-value derived from ANOVA-test

[b] p-value derived from chi-square test

[c] statistically significant difference between the two groups

**Supp Table II**: Echocardiographic findings among CardioOnco study participants stratified by cardiotoxic exposure

| | Total | Anthracyclines | Heart-relevant RT | Both | Standard risk group[a] | p-value[b] |
|---|---|---|---|---|---|---|
| n (%) | n=487 | n=260 (53%) | n=28 (6%) | n=85 (18%) | n=114 (23%) | |
| **2D LVEF mean and SD** | 59.5 ± 5.7 | **58.8 ± 5.6** **p<0.01[c]** | 59.8 ± 7.6 p=0.24[c] | **58.8 ± 5.9** **p<0.01[c]** | 61.2 ± 5.1 | **<0.01** |
| **Prevalence of abnormal LVEF % (n)[d]** | **7.2%** (35) CI (2.5% - 12.2%) | **7.3%** (19) CI (4.4% - 11.1%) | **3.6%** (1) CI (0.0% – 18.3%) | **9.4%** (8) CI (4.1% - 17.7%) | **6.1%** (7) CI (2.5% - 12.2%) | **0.68[e]** |

Abbreviations: n, number; SD, standard deviation; RT, radiotherapy; LVEF, left ventricular ejection fraction;

[a] Patients received any systemic anticancer treatment other than anthracyclines or chest-directed radiotherapy

[b] P-value received from ANOVA test

[c] T-test between different risk groups vs standard risk group

[d] Abnormal LVEF defined as <52% for males and <54% for females

[e] P-value derived from chi-square test

**Supp Table III**: Cancer history and treatment of longitudinally assessed CardioOnco study participants

| | *n = 140 (100%)* | |
|---|---|---|
| | **n** | **(%)[a]** |
| **Sex** | | |
| Female | 63 | 45% |
| Male | 77 | 55% |
| **Age at examination, Median (IQR)** | 33 (25 - 40) | |
| **Risk group** | | |
| Anthracyclines only | 80 | 57% |
| heart-relevant RT only | 10 | 7% |
| Anthracycline & heart-relevant RT only | 43 | 31% |
| Standard risk group | 7 | 5% |
| **HSCT,** yes | 13 | 9% |
| **Relapse,** yes | 18 | 13% |
| **ICCC-3 cancer diagnosis** | | |
| I Leukemias | 42 | 30% |
| II Lymphomas | 41 | 29% |
| III CNS tumors | 8 | 6% |
| Other tumors | 49 | 35% |

Abbreviations: n, number; IQR, interquartile range; HSCT, hematopoietic stem cell transplantation; RT radiotherapy; ICCC, International Classification of Childhood Cancer; CNS, central nervous system;

[a] Column percentages are given.

**Supp Table IV**: Association between risk factors and LVEF among CardioOnco Study participants retrieved from univariable linear regression

| | Unit / Reference | β[a] | 95%CI | p-value[b] |
|---|---|---|---|---|
| **Age at Study** | 1 year | **-0.05** | **(-0.11, 0.009)** | **0.09** |
| **Sex, male** | female | **-1.48** | **(-2.50, -0.46)** | **0.004** |
| **Cancer diagnoses** | | | | |
| Leukemias | Ref. | | | |
| Lymphomas | | 0.89 | (-0.47, 2.25) | 0.2 |
| Central nervous system neoplasms | | 1.22 | (-1.08, 3.52) | 0.3 |
| Other chidhood cancer diagnoses | | -0.55 | (-1.76, 0.65) | 0.37 |
| **Cumulative anthracyclines** | 100 mg/m² | **-0.81** | **(–1.15, –0.46)** | **<0.001** |
| **Heart-relevant RT** | 1 Gray | -0.008 | (-0.05, 0.03) | 0.67 |
| **Heart-relevant RT categories** | | | | |
| 0 gray | Ref. | | | |
| <15 gray | | -1.18 | (-3.52, 1.15) | 0.32 |
| ≥15 & <30 gray | | 0.08 | (-1.62, 1.78) | 0.93 |
| ≥30 | | -0.8 | (-2.73, 1.12) | 0.41 |
| **Alkylating agents[c]** | g/m2 | **-0.12** | **(-0.20, -0.04)** | **0.003** |
| **Vincristine** | mg/m2 | -0.06 | (-1.12, 0.99) | 0.91 |
| **Cisplatin** | 100 mg/m2 | **-0.32** | **(-0.67, 0.27)** | **0.07** |
| **Steroids** | g/m2 | -0.02 | (-0.18, 0.14) | 0.82 |
| **HSCT , yes** | No | **2.4** | **(0.49, 4.32)** | **0.01** |
| **Diabetes, yes** | No | **-2.72** | **(-5.88, 0.44)** | **0.09** |
| **Dyslipidemia, yes** | No | **-2.59** | **(-4.68, -0.50)** | **0.02** |
| **Hypertension, yes** | No | -0.87 | (-2.24, 0,50) | 0.21 |
| **Waist-Hip Ratio** | | | | |
| Non-obese | Ref. | | | |
| Abdominal obese | | 0.38 | (-0.81, 1.56) | 0.53 |
| **Smoking** | | | | |
| Never | Ref. | | | |
| Former | | -0.87 | (-2.45, 0.71) | 0.28 |
| Current | | -0.22 | (-1.6, 1.16) | 0.76 |

Abbreviations: HSCT, hematopoietic stem cell transplantation; RT, radiotherapy; Ref, reference;

[a] Beta coefficient is the degree of change in the LVEF value for one subgroup of patients compared to the reference subgroup.

[b] p-value retrieved from Wald test.

[c] Includes cyclophosphamid and ifosfamid.

**Supp Figure I**: Participation tree of the CardioOnco Study

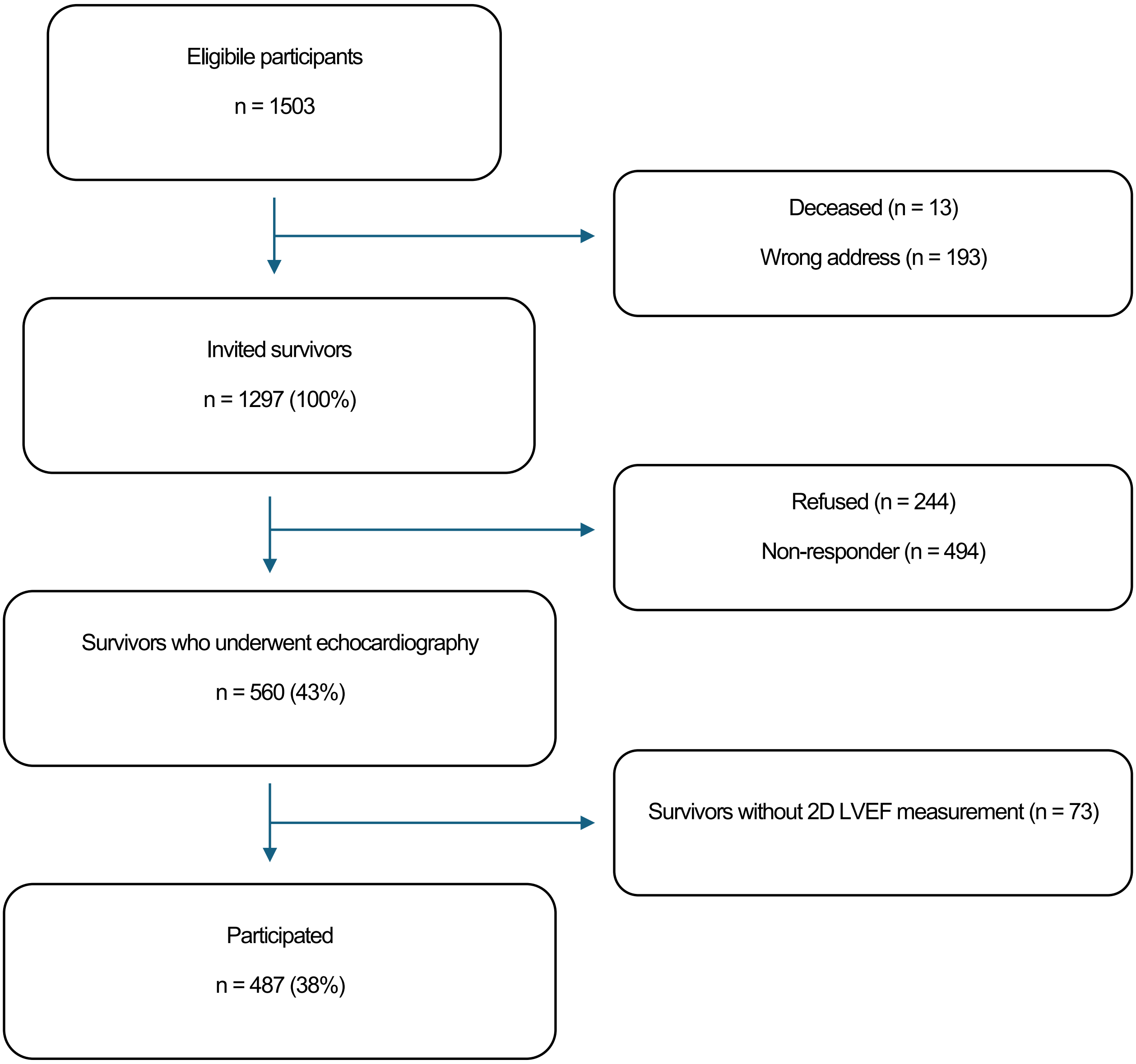

Abbreviations: n, number; LVEF, left ventricular ejection fraction;

**Detailed description of multilevel modeling**

We evaluated temporal trends using linear, quadratic, and cubic regression models, incorporating both fixed and random effects. Time (measured in years since baseline assessment) was modeled as both a fixed effect and a random slope, allowing individual variation in the rate of change in LVEF over time. Participant ID was included as a random intercept to account for repeated measurements within individuals. We compared the performance of linear, quadratic and cubic models using the likelihood ratio (LR) test, Akaike Information Criterion (AIC), and Bayesian Information Criterion (BIC). Multi-level analysis and model comparison indicated that the linear model was more appropriate than the quadratic and cubic models. The random slope and random intercept for time (years) were not statistically significant, suggesting no evidence of meaningful between-subject differences.
